# Abnormal thrombosis and neutrophil activation increases the risk of hospital-acquired sacral pressure injuries and morbidity in patients with COVID-19

**DOI:** 10.1101/2022.07.07.22277374

**Authors:** Jatin Narang, Samreen Jatana, András K. Ponti, Ryan Musich, Joshua Gallop, Angela H. Wei, Sokhna Seck, Jessica Johnson, Lynne Kokoczka, Amy S. Nowacki, Jeffrey D. McBride, Eduardo Mireles-Cabodevila, Steven Gordon, Kevin Cooper, Anthony P. Fernandez, Christine McDonald

## Abstract

Hospitalized patients have an increased risk of developing hospital-acquired sacral pressure injury (HASPI). However, it is unknown whether SARS-CoV-2 infection affects HASPI development. To explore the role of SARS-CoV-2 infection in HASPI development, we conducted a single institution, multi-hospital, retrospective study of all patients hospitalized for ≥5 days from March 1, 2020 to December 31, 2020. Patient demographics, hospitalization information, ulcer characteristics, and 30-day-related morbidity were collected for all patients with HASPIs, and intact skin was collected from HASPI borders in a patient subset. We determined the incidence, disease course, and short-term morbidity of HASPIs in COVID-19(+) patients, and characterized the skin histopathology and tissue gene signatures associated with HASPIs in COVID-19 disease. COVID-19(+) patients had a 63% increased HASPI incidence rate, HASPIs of more severe ulcer stage (OR 2.0, p<0.001), and HASPIs more likely to require debridement (OR 3.1, p=0.04) compared to COVID-19(-) patients. Furthermore, COVID-19(+) patients with HASPIs had 2.2x odds of a more severe hospitalization course compared to COVID-19(+) patients without HASPIs. HASPI skin histology from COVID-19(+) patients predominantly showed thrombotic vasculopathy, with the number of thrombosed vessels being significantly greater than HASPIs from COVID-19(-) patients. Transcriptional signatures of COVID-19(+) samples were enriched for innate immune responses, thrombosis, and neutrophil activation genes. SARS-CoV-2 viral transcripts were detected in skin tissue of COVID-19(+) patients with severe disease. Overall, our results suggest that immunologic dysregulation secondary to SARS-CoV-2 infection, including neutrophil dysfunction and abnormal thrombosis, may play a pathogenic role in development of HASPIs in patients with severe COVID-19.

**One Sentence Summary:** SARS-CoV-2-induced immune dysregulation contributes to pressure-induced sacral skin ulceration in hospitalized patients with severe COVID-19.

## Introduction

Patients with coronavirus disease 2019 (COVID-19) develop multi-organ manifestations and tissue damage secondary to both intrinsic properties of the SARS-CoV-2 virus and the resultant host immune response (*1*). The skin is among the organs affected by COVID-19, with manifestations including various localized or diffuse exanthems and retiform or acral purpura associated with thrombotic vasculopathy (*2–6*). These manifestations may cause minor adverse symptoms and exacerbate patient morbidity, but significant skin damage is rare.

Sacral ulcers are a leading cause of cutaneous-related patient morbidity in hospitalized patients (*7, 8*). The most common cause of sacral ulcers in hospitalized patients are hospital-acquired sacral pressure injuries (HASPIs), which occur secondary to unremitting pressure on the sacral and/or buttocks region (*9, 10*). Given the significant risk of HASPIs in hospitalized patients, most hospitals have developed protocols involving monitoring for early pressure-induced changes and routine off-loading measures in attempts to prevent them. Risk factors for HASPI formation include prolonged periods of immobility, decreased sensory perception, decreased mobility, increased moisture, and poor nutrition, many of which are typically present in hospitalized COVID-19 patients (*9*). Although small case series have reported the development of sacral ulcers in hospitalized COVID-19 patients, detailed analyses concerning incidence and prevalence of such ulcers, as well as risk factors associated with their development, are lacking (*11, 12*).

Despite the frequent presence of common HASPI risk factors, our clinical experience is that some COVID-19 patients acquire HASPIs that develop and progress more rapidly than usual, and which may be associated with retiform purpura (*11, 13*). Based on these observations, we hypothesized that factors inherent to SARS-CoV-2 infection contribute to formation of HASPIs in some COVID-19 patients. This study explored the pathogenesis and determined the risk and associated morbidity of HASPIs in hospitalized COVID-19 patients. In addition to characterizing the clinical features of COVID-19(+) patients with HASPIs, histologic features of sacral skin biopsies acquired from COVID-19(+) patients with HASPIs were compared to sacral skin samples from pressure ulcers (PU) in COVID-19(-) patients. Furthermore, the transcriptional landscapes of sacral skin samples from COVID-19(+) and COVID-19(-) patients were analyzed to identify SARS-CoV-2-associated contributions.

We found that COVID-19(+) patients are at an increased risk of developing HASPIs and HASPI-associated morbidity compared to COVID-19(-) patients, even when controlling for baseline risk factors for ulceration. A large percentage of sacral skin biopsies from the borders of HASPIs of COVID-19(+) patients displayed thrombotic vasculopathy (TV), with significantly more thrombosed vessels per tissue section compared to sacral skin samples from PU in COVID- 19(-) patients. Furthermore, the transcriptional profile differences in sacral biopsies from COVID- 19(+) patients with TV support enhanced abnormal thrombosis and neutrophil extracellular trap (NET)-driven inflammation in sacral skin of COVID-19(+) patients compared to sacral skin specimens from PU in COVID-19(-) patients. Finally, high levels of SARS-CoV-2 transcripts were detected in sacral skin biopsies from COVID-19(+) patients with TV, implicating a direct contribution of the virus in promotion of vascular thromboses, tissue ischemia, and eventual cutaneous ulceration.

## Results

### Risk of HASPI development and severity is increased in hospitalized COVID-19 patients

To understand the risk of HASPI development in COVID-19 disease, the incidence of sacral ulcers was compared between COVID-19(+) and COVID-19(-) patients in our study cohort (**Supplementary Figure 1**). Clinical and demographic characteristics of hospitalized patients during the study period were compared based on COVID-19 status, regardless of HASPI status. On average, COVID-19(+) patients were significantly older (p<0.001), with an increased percentage of males (p=0.003) and non-whites (p<0.001) compared to COVID-19(-) patients (**Supplementary Table 1**). COVID-19(+) patients had a significantly higher baseline risk of ulcer formation, longer length of hospitalization, higher rate of intensive care unit (ICU) admission, and a higher rate of mechanical ventilation compared to COVID-19(-) patients (**Supplementary Table 1**; p<0.001 for all 3 outcomes). Of 293 patients with HASPIs, COVID-19(+) patients had a HASPI occurrence of 7.5 per 10,000 days (49 patients; 16.7%), while HASPIs occurred in COVID-19(-) patients at 4.6 per 10,000 days (244 patients; 83.3%) (**Supplementary Table 1**). This translated to a 63% increased incidence and a 2-fold higher relative risk for HASPIs in COVID-19(+) patients compared to COVID-19(-) patients.

### COVID-19 severity is associated with HASPI risk and morbidity in relation to co-morbidities

Next, the hospitalization course of COVID-19(+) patients with and without HASPIs were compared in relation to their comorbidities (**Supplementary Table 2**). COVID-19(+) patients with HASPIs had a higher risk of ulcer formation at admission (Braden score of 14 vs. 19, p<0.001), longer hospitalization length (29 vs. 8 days, p<0.001), and more severe COVID-19 disease course, including increased requirement of mechanical ventilation or death (p<0.001) as compared to COVID-19(+) patients without HASPIs (**Supplementary Table 2**).

Multivariable regression analysis controlling for age at admission and patient co- morbidities (hypertension, chronic obstructive pulmonary disease (COPD), congestive heart failure and diabetes) was performed to assess impact of co-morbidities on HASPI development and hospitalization course (**Supplementary Table 3**). When controlling for age and comorbidities, COVID-19(+) patients who developed HASPIs were 2.2-fold more likely to have a severe COVID-19 hospitalization course compared to COVID-19(+) patients without HASPIs.

### In patients that developed HASPIs, COVID-19(+) patients are more likely to develop severe HASPIs that require debridement

Clinical and demographic characteristics in the subset of COVID-19(+) (49 patients) and COVID- 19(-) (244 patients) hospitalized patients who developed HASPIs were compared to examine the impact of COVID-19 on disease course and ulcer phenotypes (**Table 1**). Amongst patients who developed HASPIs, COVID-19(+) patients had a higher baseline risk of ulcer formation at hospital admission (Braden score of 14 vs. 18, p<0.001) and a higher frequency of requiring intubation (20.4% vs 3.3%, p<0.001) compared to COVID-19(-) patients (**Table 1**). On the other hand, gender, hospitalization length, and ICU admission rate did not differ between the two groups. A larger percentage of COVID-19(+) patients with HASPIs were black (p=0.03) (**Table 1**). Time to HASPI and HASPI size did not differ between COVID-19(+) and COVID-19(-) patients; however, COVID-19(+) patients were more likely to have a severe HASPI stage compared to COVID-19(-) patients (24.9% vs. 9.0% Stage 3 or 4 ulcer p=0.003).

**Table 1:** Clinical and Demographic characteristics of hospitalized patients who developed HASPIs.

|  |  | COVID (+) | COVID (-) | p-value<sup>a</sup> |
| --- | --- | --- | --- | --- |
| N |  | 49 | 244 | -- |
| Pressure Ulcer Risk at Admission<sup>b</sup> |  | 14 (12-16.5) | 18 (14-20) | **0.001** |
| Female Sex |  | 17 (34.7%) | 90 (36.9%) | 0.77 |
| Age (Years) |  | 66.3 (56.7-73.6) | 65.3 (55.0-73.6) | 0.31 |
| Race | White | 29 (59.2%) | 184 (75.4%) | **0.03** |
|  | Black | 19 (38.8%) | 50 (20.5%) |  |
|  | Other | 1 (2.0%) | 10 (4.1%) |  |
| Hospitalization Length (Days) |  | 29 (19.5-35) | 27 (16-41) | 0.75 |
| ICU Admission: Yes |  | 45 (91.8%) | 216 (88.5%) | 0.50 |
| Intubation: Yes |  | 10 (20.4%) | 8 (3.3%) | **<0.001** |
| Time to HASPI (Days) |  | 12 (8-20) | 14 (8-22) | 0.50 |
| HASPI Stage<sup>c</sup> | 2 | 37 | 222 | **0.003** |
|  | 3 | 7 | 17 |  |
|  | 4 | 5 | 5 |  |
| HASPI Size (cm<sup>2</sup>) |  | 12 (2.3-55.5) | 6 (1.2-29.0) | 0.06 |
| Max D-Dimer (ng/mL)<sup>d</sup> |  | 8,330<br>(3,820-18,370) | 5,490<br>(2,428-15,555) | 0.13 |
| Max Ferritin (ng/mL)<sup>d</sup> |  | 1,303<br>(909-3,582) | 813<br>(422-1,938) | **0.007** |
| Max White Blood Cell Count (K/uL)<sup>d</sup> |  | 18.5 (13.6-28.5) | 21.3 (14.9-29.7) | 0.33 |
| Max Neutrophil to Lymphocyte Ratio |  | 8.4 (4.6-12) | 9.6 (4.6-15) | 0.31 |

**Bolded** values denote statistically significant p-values.
<sup>a</sup>Based on Wilcoxon-rank sum test for continuous variable comparisons and Chi-squared test for categorical variables comparisons.
<sup>b</sup>Based on Braden risk assessment score closest to admission.
<sup>c</sup>Three COVID (+) and thirteen COVID (-) patients had unstageable HASPIs and were excluded from this analysis. P-value based on Cochran-Armitage trend test.

We also compared laboratory parameters for patients who developed HASPIs to determine if there were differences among COVID-19(+) and COVID-19(-) patients. COVID-19(+) patients had significantly higher maximum ferritin concentrations compared to COVID-19(-) patients (1,303 vs. 813 ng/mL, p=0.007). Other laboratory parameters, including maximum D-dimer concentration, maximum white blood cell (WBC) count, and neutrophil-to-lymphocyte ratio did not significantly differ between the two groups (**Table 1**).

We performed multivariable regression modeling, controlling for Braden ulcer risk at admission, hypertension, congestive heart failure, chronic obstructive pulmonary disease, diabetes mellitus, age at admission, hospitalization length, requirement for ICU admission, and requirement for intubation to assess the impact of SARS-CoV-2 infection on HASPI development and characteristics (**Table 2**). Within our cohort, COVID-19 positivity was associated with 40% increased odds of developing a HASPI (p=0.03) even after controlling for Braden ulcer risk score at admission and other variables. Additionally, COVID-19(+) patients were 2.1-fold more likely to develop a HASPI of more severe stage (p<0.001) and 3.1-fold more likely to require debridement (p=0.045) compared to COVID-19(-) patients. Time to HASPI development and wound size were not significantly associated with COVID-19 status based on multivariable regression (**Table 2**). Overall, 30-day morbidity did not differ amongst the two groups (**Supplementary Table 4**). However, a greater percentage of COVID-19(+) patients required surgical debridement compared to COVID-19(-) patients (14.3% vs. 5.7%, p=0.047; **Supplementary Table 4**).

**Table 2:** Multivariable logistic regression model assessing the impact of COVID-19 status (positive vs. negative) on HASPI development and characteristics.

| Outcome | Regression Estimate<br>(95% CI) | p-value<sup>a</sup> |
| --- | --- | --- |
| Developing a HASPI<sup>b,c</sup> | 1.4 (1.01-1.9) | **0.03** |
| Time to HASPI<sup>d</sup> | 1.3 (0.89-1.8) | 0.20 |
| Stage of HASPI<sup>e</sup> | 2.1 (1.4-3.3) | **<0.001** |
| Need for Debridement<sup>c</sup> | 3.1 (1.02-9.4) | **0.045** |
| Wound Size<sup>f</sup> | 8.1 (-0.4-16.5) | 0.06 |

**Bolded** values denote statistically significant p-values.
<sup>a</sup>P-values are representative of 5 different multivariable regression analysis all controlling for Braden ulcer risk at admission, medical comorbidities (hypertension, congestive heart failure, chronic obstructive pulmonary disease, and diabetes mellitus), age at admission, length of hospital stay, requirement for ICU admission, and requirement for intubation with COVID-19 status (positive vs. negative) as the predictor variable.
<sup>b</sup>HASPI: Hospital-acquired sacral pressure injury.
<sup>c</sup>P-value based on odds ratio from multivariable logistic regression results.
<sup>d</sup>P-value based on hazard ratio from multivariable Cox proportional hazards model.
<sup>e</sup>P-value based on odds ratio from multivariable ordinal regression.
<sup>f</sup>P-value based on parameter estimate from multivariable linear regression.

### Thrombotic vasculopathy is a prominent histopathologic feature in a significant percentage of COVID-19 patient HASPIs

The histopathological features of skin biopsies from the edges of HASPIs in COVID-19(+) patients were compared to similar tissue specimens from COVID-19(-) patients with PUs. Qualitative assessments revealed that biopsies of COVID-19(+) patients with HASPIs could be broadly categorized into 2 groups: histologic features consistent with thrombotic vasculopathy (TV; 67%) or histologic features consistent with PUs (33%); (**Figure 1; Table 3**). Whereas specimens with a TV pattern displayed diffuse fibrin thrombi within superficial and deep dermal and subcutaneous blood vessels, in addition to dermal neutrophils with occasional leukocytoclasis, specimens with a PU pattern displayed only occasional vascular thromboses with a sparse superficial dermal inflammatory infiltrate, but also displayed dermal neutrophils with occasional leukocytoclasis (**Fig. 1A and Fig.1B; Table 3**). Specimens in the COVID-19(-) PU control group typically displayed mixed dermal inflammation and dilated vascular spaces with congestion and only occasional fibrin thrombi (**Fig. 1C; Table 3**). Most specimens had an intact epidermis, but two COVID-19(-) PU control specimens had at least partial epidermal ulceration (20%). Additionally, COVID-19(-) PU specimens lacked leukocytoclasis and typically had eosinophils within the dermal inflammatory infiltrate or within vascular lumina (**Fig 1C**).

**Fig. 1.**
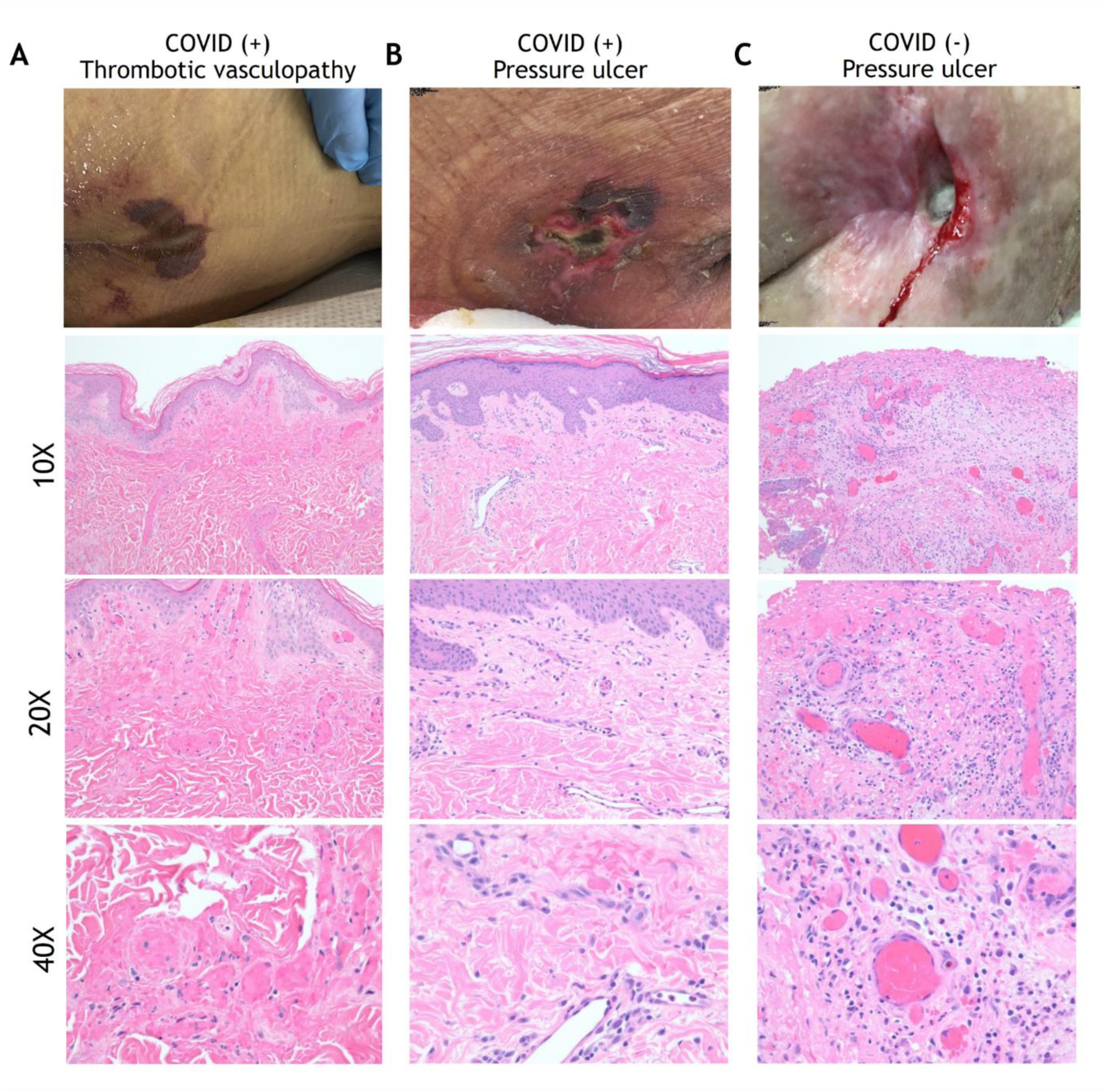
Histopathology analysis of HASPIs. (A) COVID-19-related Thrombotic Vasculopathy. A man in his 30s was intubated in the ICU due to severe COVID-19 with an acute purpuric rash on the buttocks that subsequently ulcerated. A low-power photomicrograph (100X) reveals a relatively normal and intact epidermis with diffuse fibrin thrombi involving superficial and deep dermal vessels. A higher power image (200X) further highlights diffuse fibrin thrombi and occasional inflammatory cells within the dermis. At even higher power (400X) occasional neutrophils and leukocytoclasis can be seen. (B) A woman in her 80s who developed a sacral ulcer with surrounding purpura in the setting of COVID-19 after 6 weeks of intubation. At low power (100X), a photomicrograph displays an intact epidermis with mostly patent vascular spaces in the dermis. At higher power (200X), a sparse superficial dermal inflammatory infiltrate with occasional neutrophils is noted. A 400X image reveals a rare vascular thrombosis. Other vessels display intact neutrophils within their lumens. (C) A pressure ulcer in a COVID-19(-), wheelchair-bound man in his 70s. A low-power (100X) image reveals an ulcer with superficial fibrin necrosis, mixed inflammation, and dilated vascular spaces with congestion and occasional fibrin thrombi. A higher power (200X) image reveals centrally located vessels with congestion and peripheral vessels with fibrin thrombi. At 400X magnification, a mixed inflammatory infiltrate is seen surrounding vessels that are either congested or with contain fibrin thrombi. Neutrophils are intact, without obvious leukocytoclasis. An eosinophil is present within a vascular lumen (arrow, Fig. 3C (400X)). Eosinophils were often seen in COVID-19(-)-associated ulcer specimens, either within vascular lumina or in the dermis.

**Table 3:** Clinical and pathological characteristics of biopsied patients.

|  |  | COVID (+) TV | COVID (+) PU | COVID (-) PU | p |
| --- | --- | --- | --- | --- | --- |
| N |  | 10 | 5 | 10 |  |
| Age |  | 67 (63-70) | 64 (57-78) | 55 (44-64) | 0.17 |
| % Male |  | 5 (50%) | 4 (80%) | 5 (50%) | 0.66 |
| Hospitalization Length (Days) |  | 20 (16-34) | 33 (28-66) | 15 (12-28) | 0.02 |
| Ulcer Stage | 2 | 2 (67%) | 2 (40%) | 0 (0%) | 0.04 |
|  | 3 | 0 (0%) | 1 (20%) | 1 (11%) |  |
|  | 4 | 1 (33%) | 2 (40%) | 8 (89%) |  |
| Disease Course | Hospitalization Only | 1 (10%) | 0 (0%) | 9 (100%) | <0.001 |
|  | ICU | 0 (0%) | 1 (20%) | 0 (0%) |  |
|  | Ventilation | 4 (40%) | 3 (60%) | 0 (0%) |  |
|  | Death | 5 (50%) | 1 (20%) | 0 (0%) |  |
| Max D-Dimer |  | 7,660<br>(5,465-32,580) | 5,070<br>(3,730-24,575) | 3,880<br>(3,880-3,880) | 0.39<sup>b</sup> |
| Max Absolute Neutrophil Count |  | 9.9 (4.7-17.4) | 13 (10-22) | 7.4 (3.7-15) | 0.14 |
| Pathology Features |  |  |  |  |  |
| Epidermis | Neutrophil Infiltrate | 3 (30%) | 4 (80%) | 1 (10%) | 0.031 |
| Upper Dermis | Vascular Thrombosis | 10 (100%) | 5 (100%) | 5 (50%) | 0.01 |
|  | # of Thromboses | 53 (26-99) | 12 (7-26) | 1 (0-12) | 0.0006 |
|  | Intraluminal Neutrophils | 8 (80%) | 4 (80%) | 9 (90%) | 0.99 |
|  | Leukocytoclasia | 9 (90%) | 4 (80%) | 1 (10%) | 0.001 |
| Deep Dermis | Vascular Thrombosis | 10 (100%) | 2 (40%) | 6 (60%) | 0.02 |
|  | # of Thromboses | 15 (6-20) | 0 (0-3) | 2 (0-6) | 0.001 |
|  | Intraluminal Neutrophils | 9 (90%) | 3 (60%) | 9 (90%) | 0.47 |
|  | Leukocytoclasia | 9 (90%) | 3 (60%) | 3 (30%) | 0.07 |
| Subcutis | Vascular Thrombosis | 8 (80%) | 1 (20%) | 2 (20%) | 0.20 |
|  | # of Thromboses | 4 (1-11) | 0 (0-2) | 2 (0-3) | 0.13 |
|  | Intraluminal Neutrophils | 7 (70%) | 1 (20%) | 2 (20%) | 0.32 |
|  | Leukocytoclasia | 5 (50%) | 0 (0%) | 1 (20%) | 0.29 |

<sup>a</sup>3 patients had unstageable ulcers and 1 had DTI
<sup>b</sup>Comparison between COVID(+) groups only as only 1 patient in COVID(-) had D-dimer value

The clinical and histological characteristics of biopsied patients were further analyzed in a quantitative manner (**Table 3**). Age and sex of patients with sacral tissue specimens did not differ, however average hospitalization length and severity of disease course of biopsied COVID-19(+) patients (both TV and PU) was significantly greater than in COVID(-) patients (p=0.02 and p<0.001, respectively). COVID-19(-) patients with tissue specimens available had ulcers of more severe stage compared to the two COVID-19(+) groups. Additionally, due to frequent consultation to evaluate unusual purpuric sacral rashes in COVID-19(+) patients, skin tissue specimens were taken from COVID-19(+) patients at time points closer to onset of ulceration compared to the COVID-19(-) patients (19.5 vs. 59.5 days). There were no significant differences observed in the maximum D-dimer levels and the absolute neutrophil counts between the three groups; however, not all subjects had D-dimer levels reported, especially in COVID-19(-) patients. Histologically, COVID-19(+) TV biopsies had a significantly greater number of vascular thromboses in the superficial and deep dermis, but not in the subcutis, and significantly more leukocytoclasis in the upper dermis, compared to COVID-19(+) PU and COVID-19(-) PU histolotic sections (**Table 3**).

### SARS-CoV-2-specific transcripts are elevated in COVID-19(+) TV skin tissue

The combination of clinical retiform purpura and histologic thrombotic vasculopathy in the majority of the COVID-19(+) patients with HASPIs led us to hypothesize that immunologic dysregulation due to SARS-CoV-2 infection may contribute to the pathogenesis of HASPIs in these patients. To explore this, the differentially expressed gene (DEG) profile of the tissue specimens from the TV and PU COVID-19(+) groups were compared to the COVID-19(-) PU controls. Dimensionality reduction using principal component analysis (PCA) revealed that the COVID-19(+) TV and PU groups segregated into separate clusters that were distinct from the COVID-19(-) PU control group (**Fig. 2A**).

**Fig. 2.**
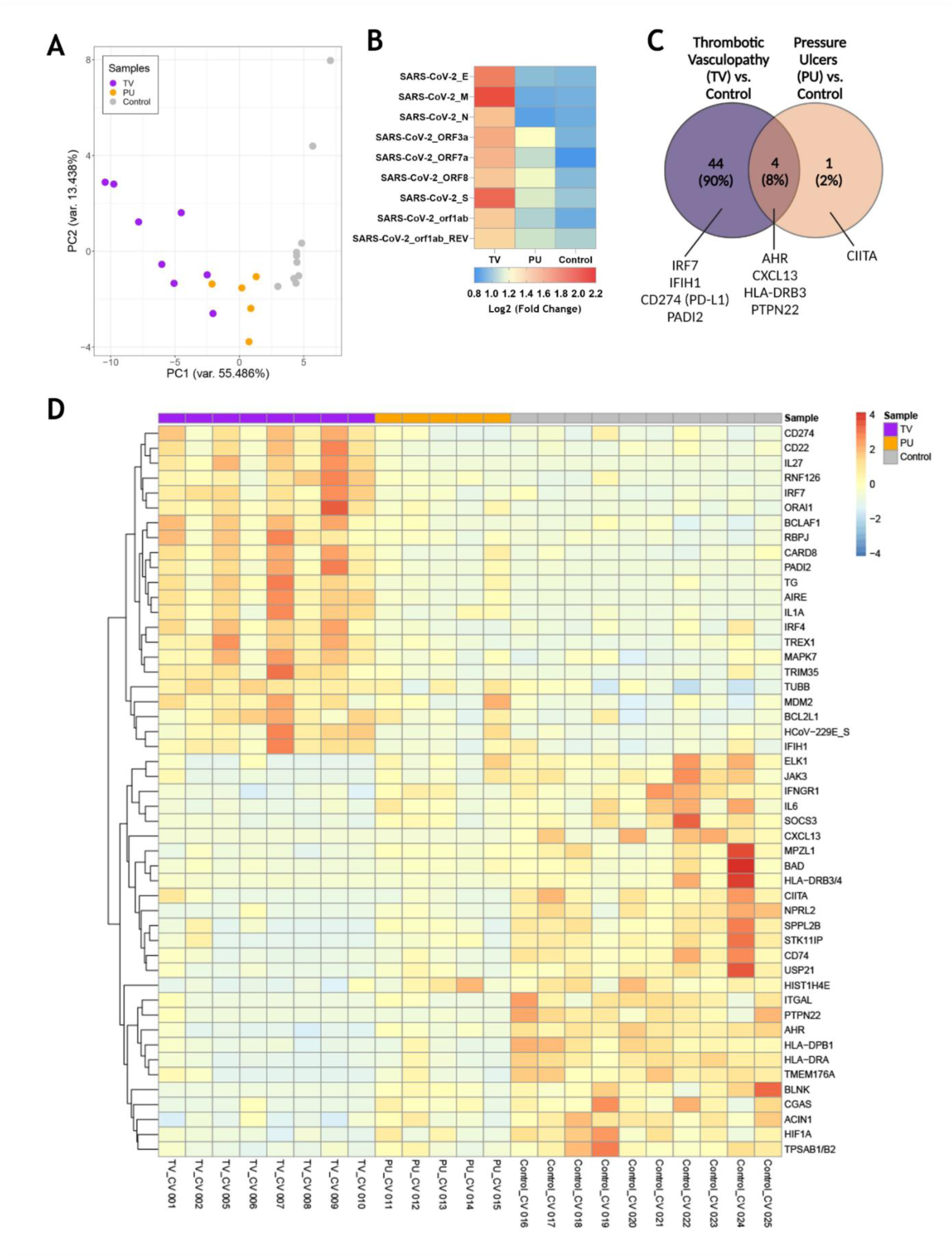
Transcriptomic analysis of sacral ulcer biopsies from COVID-19(+) patients reveals distinct signatures between TV and PU groups relative to COVID-19(-) PU control ulcers. **(A)** Principal component analysis of all patient samples using normalized counts from the differentially expressed genes with FDR adjusted *p*-value of < 0.05 indicates the distance between COVID-19(+) TV (TV), COVID-19(+) PU (PU), and COVID-19(-) PU (Control) samples. **(B)** Supervised two-dimensional Hierarchical clustering of the 49 genes whose average normalized counts were significantly different in COVID-19(+) TV and PU groups compared to COVID-19(-) PU control samples (FDR adjusted *p*-value of < 0.05). **(C)** Venn diagram showing differentially expressed genes (FDR adjusted *p*-value of < 0.05) in COVID-19(+) patients with TV and PU compared with COVID-19(-) PU control. Representative genes are indicated.

Given the three distinct transcriptional clusters, the expression of SARS-CoV-2-specific transcripts was measured to determine whether transcriptional profiles and immune responses in the skin of COVID-19(+) patients may be a direct response to viral infection or a result of systemic immune responses. Transcripts of the SARS-CoV-2 membrane glycoprotein (SARS-CoV-2_M) and SARS-CoV-2 spike glycoprotein (SARS-CoV-2_S) were pronounced in skin tissue samples from the COVID-19(+) TV group and associated with expression of several other SARS-CoV-2 viral transcripts (**Fig. 2B**). SARS-CoV-2 viral transcripts were also detected in the COVID-19(+) PU group at lower levels and were absent in the COVID-19(-) PU control group. These results suggest that presence of relatively high levels of SARS-CoV-2 viral transcripts may contribute to development of thrombotic vasculopathy in the skin tissue from the COVID-19(+) TV group.

### Genes involved in interferon-related viral responses and neutrophil extracellular trap formation are uniquely upregulated in COVID-19(+) TV skin

Among 44 DEGs that were unique to the COVID-19(+) TV group (**Fig. 2C**), genes involved in responses to viral infection, such as the interferon regulatory factor 7 (IRF7) transcription factor and interferon-induced with helicase C domain 1 (IFIH1) that encodes the cytosolic nucleic acid sensor MDA-5, were identified among the top TV-specific up-regulated genes compared to COVID-19(+) PU and COVID-19(-) PU groups. CD274 (PD-L1), which has been implicated in dysregulated neutrophil responses in severe COVID-19, and PADI2, an enzyme involved in the citrullination of arginine residues and neutrophil extracellular trap (NET) formation, were also highly expressed in the TV group compared to the other two groups (*14*).

HLA-DRB3, CXCL13, PTPN22 and AHR were identified as genes that were commonly down- regulated in both COVID-19(+) TV and COVID-19(+) PU groups compared to the COVID-19(-) PU control group. These genes are key players in antigen processing and T and B cell homeostasis. Only CIITA, a master regulator of MHC class II genes, was differentially expressed and down- regulated in the COVID-19(+) PU group compared to the COVID-19(-) PU controls. Taken together, this data suggests that innate immune responses are primarily dysregulated in COVID- 19(+) TV pathology in comparison to COVID-19(-) PUs. In contrast, antigen processing and T and B cell signaling mechanisms are potentially downregulated in both COVID-19(+) TV and PU groups.

### Hierarchical clustering of differentially-expressed genes reveals unique immunologic pathway dysregulation in COVID-19(+) TV skin compared to COVID-19(+) and (-) PU skin

To better characterize the transcriptional signatures in skin tissue from the three ulcer groups, hierarchical clustering of the top up- and down-regulated genes in each disease category was performed (**Fig. 2D**). The signatures associated with the COVID-19(+) TV and COVID-19(-) PU group were distinct from one another, supporting unique biological mechanisms of disease pathogenesis in sacral skin as a result of severe SARS-CoV-2 infection. Cumulatively, only 22 genes were similarly highly expressed in the COVID-19(+) TV, COVID-19(+) PU, and COVID- 19(-) PU groups. In comparison to the COVID-19(-) PU controls, genes highly expressed in the TV group fell into categories that included neutrophil dysfunction (CD274, PADI2), inhibition of B and T cell responses (CD22, IRF4, ORAI1), and upregulation of pro-inflammatory response pathways (CARD8, MAPK7, IRF7, IFIH1). In contrast to COVID-19(+) TV, highly expressed in the COVID-19(-) PU control group included MHC class II antigen processing (HLA-DRB3/4, HLA-DPB1, HLA-DRA, CIITA, and CD74), mast cell degranulation (TPSAB1/B2), B cell activation (CXCL13, BLNK, IL-6) and histone modification (USP21, HIST1H4E). An apoptotic gene signature was found in both COVID-19(+) TV and COVID-19(-) PU control groups but not in the COVID-19(+) PU samples, consistent with the level of tissue necrosis in these tissue specimens. However, these apoptotic gene signatures were characterized by distinct DEGs expressed in the COVID-19(+) TV (BCLAF1, BCL2L1) and COVID-19(-) PU control groups (BAD, ACIN1, HIF1A) (**Fig. 1C**). Taken together, these results indicate distinct transcriptional differences in HASPI skin of COVID-19(+) TV patients compared to COVID-19(+) PU and COVID-19(-) PU controls. Genes that were highly upregulated in COVID-19(+) TV were downregulated in both PU groups indicative of similar mechanisms of pathogenesis in PU. Upregulation of genes involved in neutrophil dysfunction and pro-inflammatory response mechanisms, specifically genes involved in response to viral infection, suggests that transcriptional differences observed in the COVID-19(+) TV group might be a direct result of SARS-CoV-2 infection and account for the unique histopathological findings of thrombotic vasculopathy (**Fig. 1A**).

### COVID-19(+) TV skin displays upregulation of unique biologic pathways compared to COVID-19(-) PU control skin

To understand the effect of infection and thrombotic vasculopathy in the COVID-19(+) TV group compared to the COVID-19(-) PU control group, the top 10 enriched Gene Ontology (GO) biological pathways in the COVID-19(+) TV and COVID-19(-) PU control groups were identified, followed by a sub-analysis to determine the significantly altered transcripts within these pathways (**Fig. 3**). Both distinct and common biological mechanisms were identified in each set of analyses (**Fig. 3A and Fig. 3B**). Top biological pathways in the COVID-19(+) TV group up-regulated in comparison to COVID-19(-) PU controls included positive regulation of pro-inflammatory cytokine production, defense response to virus, and the innate immune response pathway (**Fig. 3A**). Hematopoietic or lymphoid organ development and T cell activation involved in immune response represented top down-regulated pathways in the COVID-19(+) TV group compared to the COVID-19(-) PU control group (**Fig. 3B**). Antigen processing and presentation of exogenous peptide antigen via MHC class II biological pathway was also down-regulated in the COVID- 19(+) TV group compared to the COVID-19(-) PU control group, indicating hampered adaptive immune responses in the COVID-19(+) TV group (**Fig. 3B**). Pathway analysis did not reveal major pathways that were distinctly up- or down-regulated in the COVID-19(+) PU group compared to the COVID-19(-) PU group, suggesting similar biological mechanisms contribute to disease pathology of PU regardless of infection status.

**Fig. 3.**
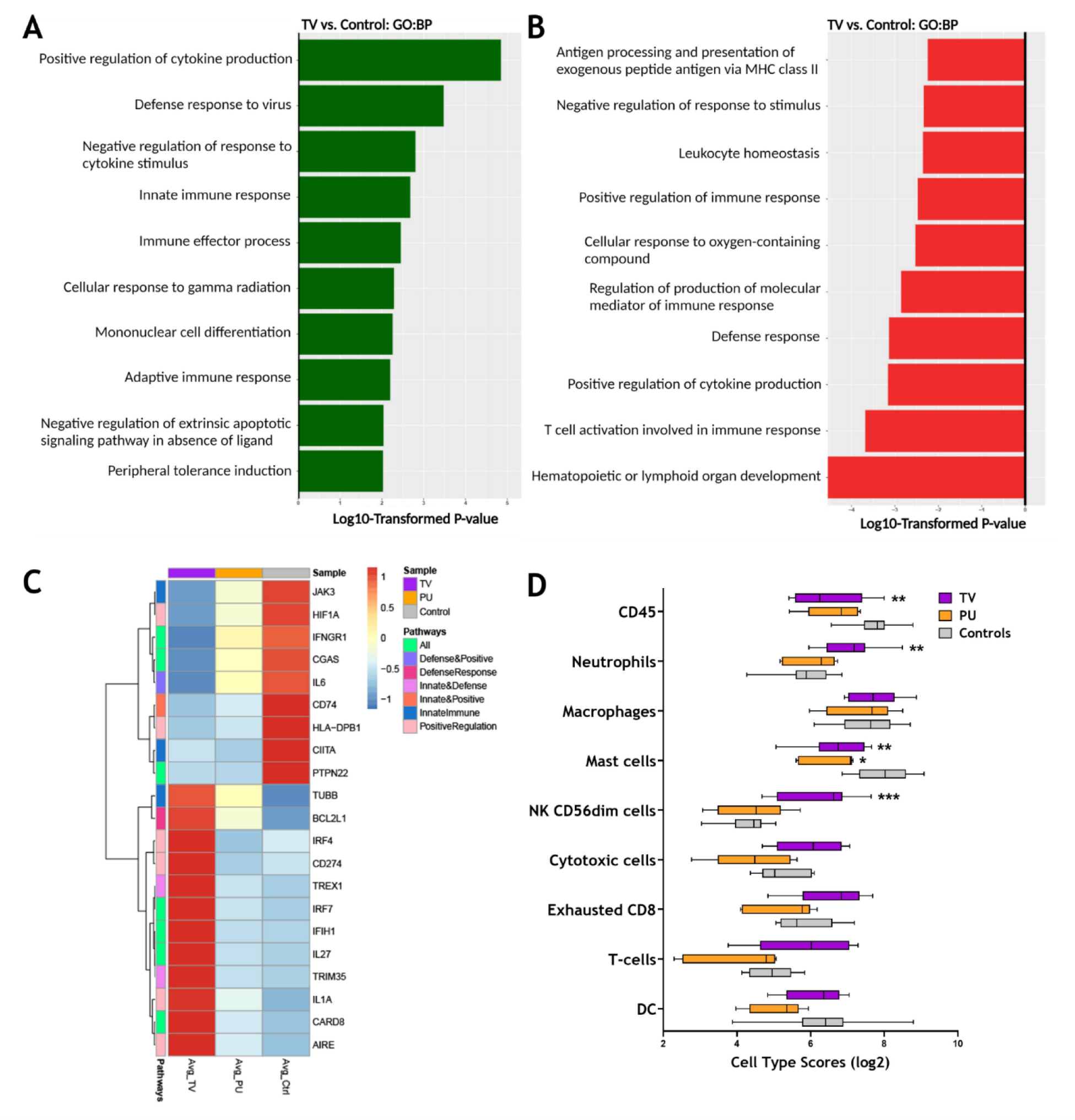
RNA expression reveals distinct immune responses in TV ulcers along with SARS-CoV-2 transcripts. **(A)** Top enriched GO biological pathways (N=10) in COVID-19(+) TV (TV) samples compared with COVID-19(-) PU (Control). **(B)** Top GO biological pathways (N=10) downregulated in TV samples relative to controls. **(C)** Clustered heatmap comparing average normalized counts of COVID-19(+) TV and COVID-19(+) PU (PU) groups with COVID-19(-) PU Control samples depicts differentially expressed genes (FDR adjusted *p*-value of <0.05) involved in positive regulation of cytokine production, defense response to virus, and innate immune response pathways. Genes involved in pathways were identified from GO biological pathways **(D)** Heatmap of SARS-CoV-2 transcripts in COVID-19(+) TV and PU groups compared to COVID-19(-) PU Controls. Data presented as means of calculated fold counts in COVID-19(+) TV and PU samples compared to COVID-19(-) PU Controls. FC, fold change. **(E)** Box and whisker plot shows the relative immune cell type scores of TV, PU, and Control samples. Cell type scores were determined using NanoString Advanced Analysis.

The analysis was expanded to uncover transcriptional signatures in the above three biological pathways up-regulated in COVID-19(+) TV specimens compared to COVID-19(-) PU control specimens (**Fig. 3C**). Select genes that were highly expressed in COVID-19(+) TV specimens compared to COVID-19(+) PU and COVID-19(-) PU control groups play a central role in regulation of innate immune responses and defense to viral infection, including CARD8, IL1A, CD274, TRIM35, IRF7, and IFIH1 (**Fig. 3C**). Both PU groups displayed similar trends in gene expression in the subset of genes that were upregulated in the COVID-19(+) TV group.

As several of the GO pathways altered in COVID-19(+) TV samples relate to regulation of immune cell populations, cell type scores were calculated to identify differences in the skin immune cell landscape in the tissue specimens from the three HASPI groups. Hematopoietic cells (CD45^+^), neutrophils, and CD56^+dim^ natural killer (NK) cells were significantly higher in the COVID-19(+) TV group compared to the other disease groups (**Fig. 3D**). The identification of a neutrophil gene signature in the COVID-19(+) TV samples is consistent with results of our DEG and GO biological pathway analyses supporting involvement of innate immune response (CD274, PADI2), as well as histologic differences in the inflammatory cell infiltrates observed in the disease groups (**Fig. 3D and Fig. 2, Table 3**). Taken together, these results demonstrate that HASPI tissue from the COVID-19(+) TV group displays upregulation of unique biologic pathways, including defense response to virus and innate immune responses, and a differential immune cell signature, compared to the COVID-19(+) PU and COVID-19(-) PU groups, suggesting differing HASPI pathogeneses. It also reinforces the histologic observations that neutrophils might be key players in COVID-19(+) TV pathology.

### Genes associated with thrombosis, PADI2-associated NET formation, and exacerbated inflammatory responses are specifically up-regulated in COVID-19(+) TV skin

Neutrophils have been implicated as key players in mechanisms contributing to immunothrombosis in COVID-19 disease (*15–17*). Given this and our above results, gene-set enrichment analysis (GSEA) was utilized to more completely characterize transcriptional signatures associated with thrombosis, immune cell chemotaxis, and neutrophil-specific biological functions in our HASPI groups. First, examined were genes involved in thrombosis and complement genes that may contribute to enhanced thrombosis. Notably, CXCL10, a cytokine that is implicated among the three key players contributing to COVID-19 cytokine storm and increased fatal COVID-19 outcomes (*18*), was highly expressed in the COVID-19(+) TV group compared to the other disease groups. Additionally, genes associated with thrombotic mechanisms, including MEFV, STAT5B, CD55, CRP, CTLA4, TLR4, CCR1, CFP, C4A/B and IL23R, were only upregulated in the COVID-19(+) TV group (**Fig. 4A**). Alternatively, complement activation genes were similarly expressed in all three ulcer groups (**Fig. 4A**). These findings suggest that COVID- 19(+) TV HASPIs may have abnormal thrombosis without enhanced complement activation due to upregulation of genes that mediate thrombosis.

**Fig. 4.**
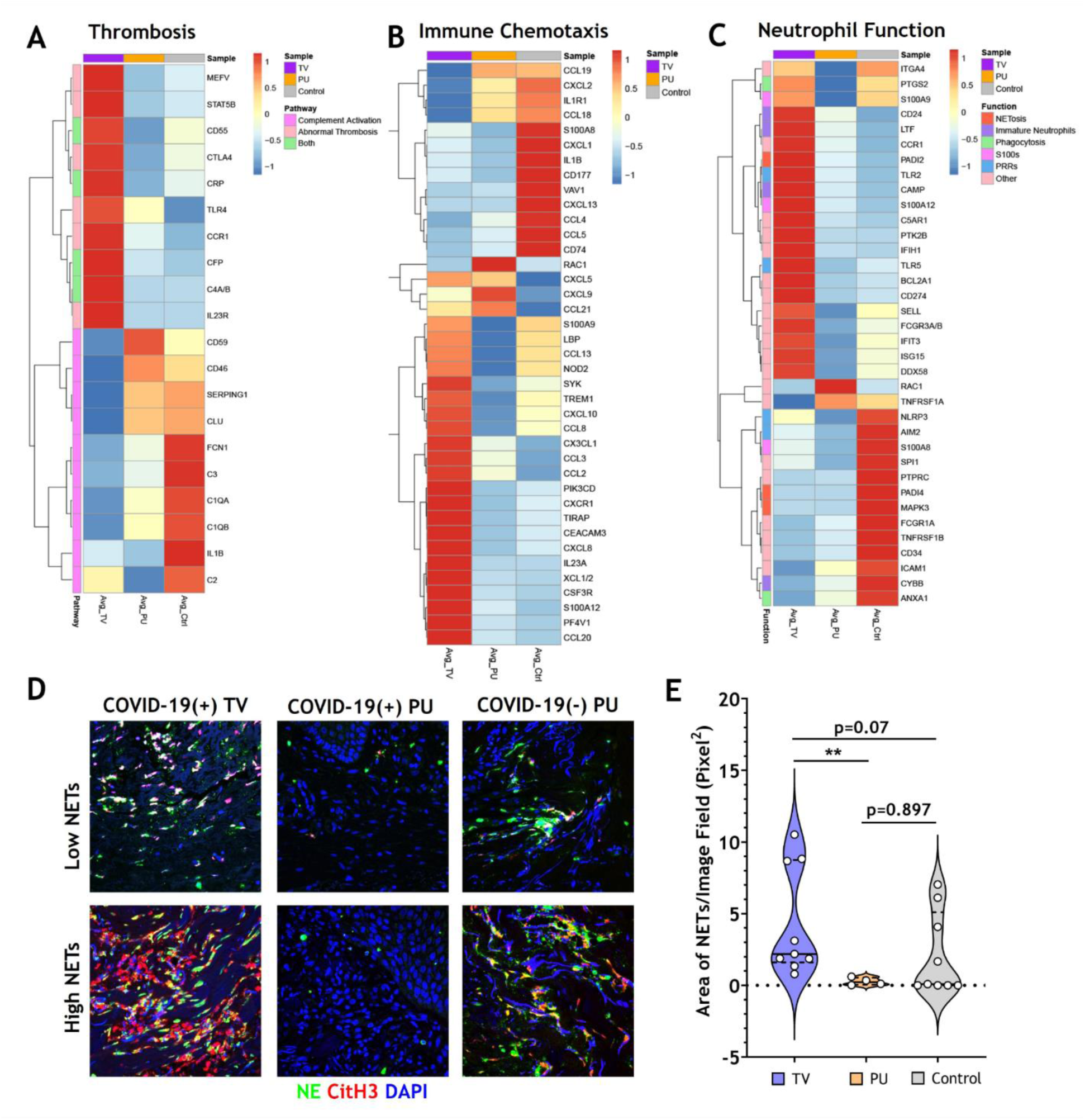
COVID-19 thrombotic vasculopathy is characterized by abnormal thrombosis and neutrophil extracellular trap formation. **(A)** Clustered heatmap shows the average normalized counts of genes involved in thrombosis and complement activation for COVID-19(+) TV (TV) and COVID-19(+) PU (PU) samples compared to COVID-19(-) PU (Control). Pathway specific genes were selected based on GO biological pathways. **(B)** Clustered heatmap illustrates the average normalized counts of genes involved in immune cell chemotaxis and extravasation for COVID-19(+)TV and PU samples compared to COVID-19(-) PU Controls. Genes were selected based on GO biological pathways **(C)** Neutrophil specific genes arranged by function (NETosis, Immature Neutrophils, Phagocytosis, S100s, PRRs, or Other) represented in a clustered heatmap comparing the average normalized counts of DEGs in COVID-19(+) TV and PU groups relative to COVID-19(-) PU Controls. **(D)** Immunofluorescence staining of NETs in skin tissue sections. Markers of NETs used include NE (green), CitH3 (red) and nuclei (blue). Images acquired using confocal microscopy, 40X oil objective **(E)** Quantification of tissue NETs in the skin using Image J analysis.

Next, the dataset was surveyed for all genes that are involved in immune cell chemotaxis and extravasation (**Fig. 4B**). In both COVID-19(+) TV and COVID-19(-) PU control groups a gene expression signature associated with neutrophil activation and chemotaxis was expressed; however, these signatures were characterized by unique subsets of genes in COVID-19(+) TV (TREM1, CXCR1, CXCL8, and S100A12) and COVID-19(-) PU control (CXCL1, CXCL2, and CD177) groups (**Fig. 4B**). A small subset of genes involved in neutrophil and T cell activation (CXCL5, CXCL9, CCL21) were expressed in both COVID-19(+) TV and COVID-19(-) PU groups. Additionally, genes associated with antimicrobial defense mechanisms, including alarmins (S100A9, S100A12) and pattern recognition receptors (LBP, and NOD2), were up-regulated in the COVID-19(+) TV group compared to the COVID-19(-) PU control group, while suppressed in the COVID-19(+) PU group. While some genes involved in monocyte and lymphocyte migration and function were expressed in the COVID-19(+) TV group (CCL13, CCL8, and CCL20), a greater number of these genes were expressed in the COVID-19(-) PU control group (CCL19, CCL18, CXCL13, CCL4, CCL5, CXCL9). Only two genes (RAC1, CXCL9) were upregulated in COVID-19(+) PU compared to the other two disease groups. These genes are involved in the control of cell growth and lymphocyte chemoattraction, respectively. These findings support the histologic findings of different immune cell populations present in the HASPI groups. Taken together, these results demonstrate that while neutrophil activation and dysfunction may play a role in HASPI skin of both COVID-19(+) TV and COVID-19(-) PU groups, abnormal thrombosis and exacerbated inflammatory responses appear to be unique to HASPI skin in the COVID-19(+) TV group.

### NET formation and upregulation of genes associated with the immature neutrophil subset are significantly elevated in COVID-19(+) TV skin

Schulte-Schrepping et al. utilized single-cell RNA sequencing of whole blood and peripheral blood mononuclear cells from COVID-19(+) patients to study the role of mature and immature neutrophil subsets in COVID-19 disease pathogenesis (*14*). We utilized this published dataset of neutrophil function-related genes that contribute to systemic inflammation in COVID-19 disease and compared them to the transcriptional signatures in our HASPI skin tissue dataset. Genes contributing to dysregulated NET formation were highly expressed in both COVID-19(+) TV and COVID-19(-) PU groups, but not in the COVID-19(+) PU group (**Fig. 4C**). However, genes related to distinct mechanisms of NET induction were differentially expressed in the COVID-19(+) TV and COVID-19(-) PU control groups, with COVID-19(+) TV samples expressing PADI2 versus PADI4 expression in COVID-19(-) PU controls.

Immature neutrophils, or low-density granulocytes, have been shown to undergo uncontrolled NET formation in several autoinflammatory and autoimmune diseases, contributing to disease pathology (*19*). Genes associated with the immature neutrophil subset (CD24, LTF, CAMP) were up-regulated in COVID-19(+) TV skin, but not in COVID-19(+) PU or COVID-19(-) PU skin tissue (**Fig. 4C**). Several other neutrophil function-related genes were also uniquely expressed in the COVID-19(+) TV group, including those related to antiviral response (IFIT3, DDX58, PTGS2, and IFIH1) and Toll-like receptor (TLR)-mediated signaling (TLR2, TLR5). Finally, CD274 (PD-L1), a known inhibitor of T cell activation, was highly expressed in COVID- 19(+) TV skin tissue compared to the other 2 groups. These results reveal that while certain neutrophil chemotactic genes are expressed in both COVID-19(+) TV and COVID-19(-) PU control groups, expression of genes associated with the immature neutrophil subset is a distinct characteristic of the COVID-19(+) TV group, suggesting this neutrophil subset might contribute to enhanced vascular thrombosis at the local skin tissue level.

Finally, NETs in skin tissue from all three HASPI groups were visualized by immunofluorescent staining. Rare NETs were detected in the COVID-19(+) PU group (**Fig. 4D**), which is consistent with the low NET-specific DEGs detected in this group (**Fig. 4C**). This is in contrast to the visualization of NETs mainly in the dermis in all of the COVID-19(+) TV samples, which ranged from relatively low to extensive NETosis (**Fig. 4D**). NETs were also variably detected in COVID-19(-) PU control tissue specimens, with five of nine COVID-19(-) PU skin specimens (∼55%) demonstrating a lack of detectable NETs (**Fig. 4D**). This was supported by image analysis to quantify the area of NETs within the tissue, which demonstrated the median total NET area in the COVID-19(+) TV group was modestly elevated compared to COVID-19(-) PU controls (p=0.07) (**Fig. 4E**). Total area of NETs in the COVID-19(+) TV tissue was significantly higher than in the COVID-19(+) PU group (p<0.01), indicating a potential role for neutrophils in abnormal thrombosis-mediated biological mechanisms. These results demonstrate that while NETs may contribute to pathology in both COVID-19(+) TV and COVID-19(-) PU groups, mechanisms of NET induction (PADI2 vs. PADI4) and neutrophil subsets that potentially contribute to disease pathology (immature vs. mature neutrophil subsets) differ, possibly due to local SARS-CoV-2 infection.

## Discussion

Although the hallmark of COVID-19 disease is acute respiratory distress syndrome and respiratory failure, manifestations are known to occur in many organ systems (*20*). Inflammation related to COVID-19 can cause transient or permanent organ damage, adding considerable morbidity to patients who survive their COVID-19 disease course. Numerous cutaneous eruptions have been described in the setting of COVID-19, but the peer-reviewed literature has focused mainly on outbreaks that resolve without significant tissue damage. Alternatively, HASPIs represent localized areas of significant and sometimes permanent skin damage. HASPIs occur mainly in hospitalized patients and are associated with substantial patient morbidity and healthcare costs, with estimates ranging from $10 billion to >$26 billion annually (*21*). Given the significant impact of HASPIs and requirement for hospitalization in a substantial percentage of patients with COVID- 19, we explored the incidence of HASPIs and risk factors for their development in a large cohort at a single tertiary care, multi-hospital institution.

This study revealed a number of important findings surrounding HASPIs in COVID-19(+) patients. Overall, COVID-19(+) patients had a 63% increased incidence of developing a HASPI compared to COVID-19(-) patients (**Supplementary Table 1**). Additionally, COVID-19(+) hospitalized patients have a 40% increased odds of developing HASPIs compared to COVID-19(-) patients, even when controlling for baseline ulceration risk at admission, ICU admission, and other variables (**Table 2**). Although the overall etiology of HASPIs in COVID-19 patients is likely multifactorial, there are numerous possible explanations for the increased risk we found in these patients. One obvious possibility is that COVID-19(+) patients have increased risk of ulceration at time of admission. However, risk of HASPIs in COVID-19(+) patients persisted compared to COVID-19(-) patients when controlling for risk of ulceration in a multivariable model. Another possibility is that meticulous routine care for these patients was compromised to protect caregivers from excess exposure to COVID-19(+) patients. However, protocols for frequent skin checks for HASPIs were not changed at our institution during the COVID-19 pandemic. This makes lack of adequate care an unlikely cause of increased HASPIs in our COVID-19(+) patient population and suggests other factors contribute to this increased risk, potentially including factors inherently related to SARS-CoV-2 infection.

This study identified that COVID-19(+) patients are at risk of more severe HASPIs and more often require surgical debridement compared to COVID-19(-) patients (**Table 1 and Table 2**). In particular, the finding that almost 25% of the COVID-19(+) HASPIs were Stage 3 or Stage 4 (compared to <10% in the COVID-19(-) cohort) is important, as these stages of pressure injuries are considered hospital ‘never events’. These ‘never events’ were defined by the National Quality Forum as events that are measurable and preventable, and that may lead to severe patient morbidity and mortality. The increased incidence of these severe ulcerations, despite continuing standard clinical monitoring protocols during COVID-19 to prevent them, is concerning.

Importantly, when controlling for age, co-morbidities, and other variables COVID-19(+) patients with HASPIs were more likely to have a severe hospitalization course compared to COVID-19(+) patients without HASPI. These results support that development of a HASPI is a poor prognostic sign in the setting of COVID-19. However, it is unclear whether a severe hospitalization course increases risk of HASPI development, or vice versa. Regardless, this finding aligns with the hypothesis that HASPIs in COVID-19(+) patients likely arise as a result of both intrinsic and extrinsic disease factors.

A comparison of laboratory parameters revealed COVID-19(+) HASPI patients had significantly higher maximum levels of serum ferritin compared to COVID-19(-) HASPI patients (**Table 1**). Hyperferritinemia has been associated with poor prognosis in COVID-19 patients and has been implicated to have a pathogenic role in cytokine storm (*22*). In the setting of an inflammatory state, ferritin has also been hypothesized to be a marker of cellular damage. Thus, COVID-19-associated inflammation may result in more significant cellular damage compared to other medical conditions leading to hospitalization, resulting in an overall catabolic state that promotes HASPI ulceration.

Surprisingly, maximum D-dimer levels were not significantly higher in COVID-19(+) patients with HASPIs compared to COVID-19(-) patients with HASPIs in our cohort. However, a high percentage of COVID-19(-) patients had no D-dimer levels drawn, suggesting a selection bias towards evaluating D-dimer levels in those patients who may have had clinical features supporting a thrombotic state (**Table 1**). Elevated D-dimer levels strongly correlate with an increased risk of thrombotic events in COVID-19 patients and, importantly, all COVID-19(+) HASPI patients had elevated D-dimer levels (*23*). Factors other than elevated D-dimer levels, such as elevated expression of thrombotic genes or heightened triggering of complement pathway activation, may also contribute to thrombotic pathology in COVID-19 patients.

SARS-CoV-2-induced thrombotic events in the skin microcirculation represent another possible contributor to increased HASPI risk in COVID-19 patients. Thrombotic events in the skin and other organs of patients with COVID-19 are well-described (*3, 4*). Although these events typically do not appear to result in significant skin damage, they may promote eventual ulceration in areas like the sacrum that are additionally affected by pressure. In support of this, we have previously seen and reported several patients with severe COVID-19 who developed unusual retiform purpura on the buttocks associated with histologic thrombotic vasculopathy and subsequent development of sacral ulcers (*11, 13*).

In our cohort, 25 patients had skin specimens of their HASPIs available, 15 of which were biopsies performed on COVID-19(+) patients (TV and PU). We compared the histologic, cellular and molecular characteristics of these specimens in detail to each other and to those in COVID- 19(-) PU control skin tissue. Significantly higher numbers of vascular thromboses and leukocytoclasis were observed in skin tissue from the COVID-19(+) TV group compared to the other groups, supporting the role of thrombotic vasculopathy as a potential contributor to subsequent ulceration in some COVID-19(+) patients.

Interestingly, vascular thromboses were detected occasionally in our COVID-19(-) PU control specimens. In fact, vascular thromboses have been described in skin biopsies of HASPIs in COVID-19(-) patients, and vascular occlusion has long been hypothesized to play an important pathogenic role in the development of PUs (*24, 25*). However, vascular thromboses were far more numerous and prominent in COVID-19(+) TV tissue compared to what we found in COVID-19(-) PU skin and compared to what has been described in prior histologic studies of PUs, suggesting additional pathogenic contributers to thrombotic vasculopathy in the COVID-19(+) TV group, such as factors inherent to COVID-19 (*15, 26, 27*). Additionally, some prior histologic studies of PUs describe vascular thromboses underlying ulcerated skin, suggesting the possibility some represent secondary phenomena related to inflammation in areas of ulcer, whereas our COVID- 19(+) TV tissue contained diffuse vascular thromboses in skin with intact epidermis, prior to ulceration (*28, 29*).

An increased presence of SARS-CoV-2-associated transcripts were detected in COVID- 19(+) TV skin tissue compared to the other groups, suggesting a potential role for the virus in promoting cutaneous thromboses. Prior work supports a role for SARS-CoV-2 viral spike protein in precipitating cutaneous vascular thromboses in COVID-19 patients. Previous histologic studies demonstrated that SARS-CoV-2 viral spike protein localizes to ACE2+ endothelial cells in cutaneous microvessels in the dermis and subcutis, as well as vessels surrounding sweat glands, and this localization has been suggested to precipitate vascular thromboses (*30, 31*). ACE2 destruction increases necrotic cell death and promotes vasoconstriction, which can worsen ischemia in areas persistently under pressure. Furthermore, hypoxia induced during endothelial cell damage further compounds a hypercoagulable state. Thus, localization of SARS-CoV-2 spike protein in endothelial cells of cutaneous vessels may cause endothelial cell damage that increases the likelihood of triggering vascular thromboses and alternative complement and coagulation pathway activation (*32*).

Although the majority of skin specimens from COVID-19(+) patients showed a TV pattern, one-third showed a PU histologic pattern with far less vascular thromboses compared to TV skin tissue. Given COVID-19(+) PU skin had less SARS-CoV-2 spike transcripts compared to COVID- 19(+) TV skin, a threshold amount of SARS-CoV-2 spike protein may be needed to contribute to the development of vascular thromboses with or without pressure-induced cutaneous changes. Additionally, although not statistically significant, mean maximum D-dimer levels were higher in the COVID-19(+) TV group compared to the COVID-19(+) PU group. This raises the possibility that there may also be a threshold D-dimer level, above which there is increased risk of vascular thrombosis, in cutaneous vessels under pressure.

Transcriptional analyses on skin specimens from the sites of sacral ulcer formation evaluated the inflammatory landscape in skin tissue during SARS-CoV-2 infection in comparison to sacral ulcers obtained from uninfected patients. This type of analysis has not been previously performed. Our results showed that COVID-19(+) TV skin is associated with up-regulation of genes related to innate immune responses and viral defense responses, and down-regulation of T-cell mediated immune responses (**Fig. 3A and Fig. 3B**). Importantly, biological pathways and DEGs upregulated in COVID-19(+) TV skin were mainly categorized as genes that would potentially contribute to abnormal thromboses and immunothrombosis, consistent with our histopathology results (**Fig. 4**). Analysis of DEGs showed that while interferon response genes and neutrophil-associated genes were highly expressed in COVID-19(+) TV skin, genes associated with MHC class II antigen presentation were down-regulated, indicative of stunted adaptive immune responses in COVID-19(+) TV (**Fig. 2C**). Consistent with results of histopathological observations (**Table 3**), we also observed that genes associated with mechanisms of dysregulated immature neutrophil responses were highly expressed in COVID-19(+) TV skin compared to COVID-19(-) PU skin (**Fig. 4B and Fig. 4C**). Given the known association between SARS-CoV-2 infection and immature neutrophil responses, this result further implicates SARS-CoV-2 infection itself with augmentation of ischemia in these areas and contribution to eventual tissue necrosis and ulceration. Alternatively, genes related to complement activation were upregulated in all disease groups, indicating that the alternative complement activation pathway was triggered in infected as well as uninfected skin tissue. Overall, our results suggest SARS-CoV-2 infection may amplify ischemia and eventual tissue necrosis and ulceration in skin under pressure by increasing thrombosis and neutrophil dysregulation, and not complement activation.

Recently, Yang et al (*33*) examined the importance of CXCL10 plasma levels (interferon γ-inducible protein 10; IP-10) in COVID-19 disease severity and progression of COVID-19 disease. They reported that 5 cytokines, including CXCL10, were associated with fatal outcomes during infection (*33*). Several other studies have also shown the prognostic value of CXCL10 in determining COVID-19 disease course (*34, 35*). CXCL10 was found to be up-regulated in COVID-19(+) TV skin in comparison to COVID-19(+) PU and COVID-19(-) PU skin in our study (**Fig. 4A**). These results, in addition to higher levels of SARS-CoV-2 transcripts in COVID-19(+) TV skin, suggest COVID-19 severity may be important for precipitating cutaneous vasculopathy in skin under pressure, which would further support a threshold is needed for SARS-CoV-2- associated contributions to cutaneous thrombosis.

Cytokines, such as IL1A and IL27, which have been shown to be higher in patients with more severe COVID-19 disease (*36–38*), were highly expressed in COVID-19(+) TV skin in comparison to the other groups. We also found that alarmins (S100A9, S100A12) and pattern-recognition receptors (PRRs) including TLR2, TLR4, TLR5, IFIH1, and NOD2 had higher expression in COVID-19(+) TV skin. It has been shown that SARS-CoV-2 spike protein binds TLR4 to increase the cell surface expression of ACE2 and facilitate viral entry (*39, 40*) exacerbating innate immune responses during infection. Expression of SARS-CoV-2-specific genes and higher transcript levels of PRRs in COVID-19(+) TV skin could potentially explain the enhanced innate immune responses observed with examination of GO biological pathways, and a contribution of neutrophil dysfunction in promoting cutaneous vascular thromboses.

Cell type scoring demonstrated that neutrophils were enriched in COVID-19(+) TV skin compared to COVID-19(+) PU and COVID-19(-) PU skin. While we also observed changes in CD56^+dim^ NK cell and mast cell signatures, we focused our subsequent efforts on specifically understanding the role of neutrophils in the ulcer phenotypes with and without SARS-CoV-2 infection since neutrophils have been shown to play an important role in COVID-19 disease pathogenesis, particularly in relation to thromboses and multi-organ damage (*17, 41, 42*). A study of dysregulated myeloid cell responses in COVID-19 revealed that presence of CD274 and ARG1 expressing dysfunctional mature neutrophils contributed to severe COVID-19 disease (*14*). In our dataset, the expression of CD274 was significantly higher in TV skin compared to skin in the other groups. Given that T-cell mediated immune responses were down-regulated in COVID-19(+) TV skin, it is possible that presence of CD274^high^ (PD-L1^+^) mature neutrophils in TV skin tissue suppress T cell responses due to the inhibitory interaction between PD-L1 and PD-1, resulting in stunted adaptive immune responses. In support of this, mature CD274 (PD-L1^+^) neutrophils have been shown to exert suppressive effects on T cell function in various other diseases like cancer and HIV infection (*43, 44*).

The presence of NETs in skin tissue was directed evaluated to understand whether exacerbated NET formation contributed to inflammation in the skin. Results showed that NETs were significantly higher in COVID-19(+) TV vs. PU skin, indicating that neutrophils and NETs potentially contribute to cutaneous thrombotic vasculopathy during severe COVID-19 infection. While NETs were also observed in COVID-19(-) PU (controls), the baseline levels in 5 patients were nearly zero, indicating that exacerbated NET formation might be a consistent feature of TV in COVID-19(+) patients.

Our study has several advantages and limitations. Advantages include the establishment of a comprehensive COVID-19 registry at our institution that allowed identification of all COVID- 19(+) patients admitted to our institution, as well as a relatively large cohort utilized for analysis. Additionally, the standard protocol to photograph ulcers allowed objective confirmation that patients were accurately labeled as having HASPIs. More importantly, our results correlate clinical findings with histopathologic and molecular results. The comparison of COVID-19(+) patients with HASPIs to both COVID-19(+) patients without HASPIs and COVID-19(-) patients with HASPIs has not previously been done, especially with quantification of genes in sacral skin under pressure in all 3 groups. We were also able to corroborate the contribution of NETs to local tissue inflammation indicated by our transcript analyses using immunofluorescence imaging.

Limitations include the retrospective nature of our study and possibility that electronic medical records did not always contain accurate documentation of data points (i.e. date of ulcer onset). Additionally, laboratory data points and skin tissue specimens were not always captured on similar days during hospitalizations or in all patients to allow precise comparison. We were unable to utilize fresh biopsies for transcript-level analyses and corroborate with systemic markers due to the unavailability of these resources in severely ill patients during the initial months of the outbreak. Therefore, the transcriptional analyses were limited to bulk tissue digests of paraffin-embedded tissue, precluding single cell transcriptomic analyses. Furthermore, as this study consists of a cohort from a single institution, our results are not necessarily generalizable. This study also focused specifically on the role of neutrophils and NETs in disease pathology; however, future studies will explore the role of other cells types that were significantly altered in the three disease groups.

In conclusion, we show that COVID-19(+) patients are at an increased risk of HASPIs compared to COVID-19(-) patients, and COVID-19(+) patients with HASPIs develop ulcerations of more severe stage that more often require debridement than COVID-19(-) patients. Furthermore, we show that thrombotic vasculopathy was prominent in sacral skin of COVID-19(+) patients with HASPIs compared to controls, and TV tissue displayed increased expression of SARS-CoV-2-specific transcripts and DEGs known to contribute to abnormal thrombosis and ischemia. Overall, our results implicate SARS-CoV-2 infection as a contributing factor to HASPIs in at least some COVID-19 patients.

Unlike other reports of skin manifestations in COVID-19 patients, our overall findings support the skin is yet another organ system that may sustain significant damage secondary to COVID-19. Recognizing risk of HASPIs and the potential role of SARS-CoV-2 infection in COVID-19 patients is important, as they may contribute to significant future morbidity, healthcare expenditure, and even mortality in patients who recover from COVID-19. Further research is needed to uncover detailed pathogenic factors leading to increased HASPI risk in COVID-19(+) patients, as well as how to optimally prevent them from occurring. For now, healthcare workers should be especially attentive to potential early signs of HASPI development in patients hospitalized with COVID-19, especially those with serious disease, and be meticulous with currently proven preventative clinical interventions.

## Materials and Methods

### Patient Identification

Our study protocol was approved by the Cleveland Clinic Institutional Review Board. Patients with HASPIs were identified as having been hospitalized for greater than 5 days within our institution between March 1 and December 31, 2020, with an ICD-10 diagnosis of sacral ulcer or photographic documentation of sacral ulceration at some point during hospitalization (**Supplementary Table 5**). We defined a sacral ulcer as any new onset ulceration involving the sacral and/or buttocks areas acquired during an inpatient hospital admission at one of our facilities within the above time period. The decision to utilize an image search was made because nursing staff at our institution are instructed to image all potential HASPIs for documentation and clinical improvement purposes, and because photographs provide objective proof of ulceration. Based on our search criteria, we assumed all sacral ulcers that developed during hospitalization represented HASPIs unless otherwise stated in clinical notes and will refer to them as such from this point forward.

Patients that did not undergo a polymerase chain reaction (PCR)-based COVID-19 test at admission or during hospitalization, those hospitalized for <5 days, and those with sacral ulcers already present at hospital admission were excluded from our study. Additionally, stage 1 ulcer patients were excluded as these types of ulcers are difficult to reliably detect and are not routinely documented by our wound care teams. Some patients were hospitalized more than once during our defined study period. However, only the hospitalization associated with an initial development of a HASPI was included for our study purposes.

Among the 143,223 patients hospitalized at our institution during our defined study period (March 2020-December 2020), 58,766 patients met our inclusion/exclusion criteria and comprised our study cohort (**Supplementary Fig. 1**). These patients were hospitalized for a total 591,763 days. Of our 58,766 patients, 5,531 (9.4%) were COVID-19(+) patients (total hospitalization days = 64,980; average 11.8 days) and 53,528 (89.6%) were COVID-19(-) patients (total hospitalization days=526,783; average 9.8 days). A total of 3,352 patients had a diagnosis of sacral ulcer during their hospitalization with or without an image of their sacral region in their electronic medical records. Of these patients, 2,992 had their sacral ulcers documented prior to or upon admission and 67 were seen at an outside hospital and were excluded from our study. Thus, our final study cohort consisted of 293 patients with HASPIs (**Supplementary Fig. 1**).

### Patient and Ulcer Characteristics

For all patients who met our inclusion/exclusion criteria, we thoroughly reviewed electronic medical records to obtain detailed clinical information. Clinical data acquired included demographic information (age, gender, race), HASPI ulceration risk (based on Braden risk assessment at hospital admission), presence of specific medical comorbidities (obesity, hypertension, diabetes mellitus, and chronic obstructive pulmonary disease (COPD)) index hospitalization information (COVID-19 status, length of stay, intensive care unit (ICU) admission, intubation/mechanical ventilation requirements), laboratory results (including maximum D-dimer concentration, maximum ferritin concentration, and maximum white blood cell count (WBC)), HASPI characteristics (time from admission to ulcer formation, ulcer stage, and ulcer size), and 30-day HASPI-related morbidity information (defined as either need for surgical debridement or subsequent ulcer-related infection within 30-days of discharge).

The Braden risk assessment score is a validated clinical tool calculated by nursing staff throughout our institution as part of routine patient skin assessments at admission and every 12 hours thereafter. This standardized risk scale assigns numerical ratings (ranging from 1-4) to six domains, including sensory perception, moisture, activity, mobility, nutrition, and friction/shear forces (*45*). The sum of these domains is used to risk stratify patients for HASPI formation; a lower score is correlated with increased risk of HASPI development (*46*).

The primary outcome of our study was development of HASPI during the indexed hospitalization. Our secondary outcome was severity of hospitalization course, which was an ordinal categorical variable defined as either (1) hospitalization only, (2) hospitalization requiring ICU admission, (3) hospitalization requiring ICU admission and need for mechanical ventilation, and (4) hospitalization leading to death.

COVID-19 status was determined by nasal swab and PCR-based diagnostic testing. If a patient tested positive for COVID-19 at least once during the index hospitalization, they were classified as COVID-19(+). Time to ulcer formation was measured in days from admission to first mention and/or photographic documentation of HAPSI in the patient’s EMR, whichever occurred first. Ulcer size was defined as the largest recorded area (measured in cm^2^) of sacral ulceration during the index hospitalization. Ulcer stage was defined as most severe stage of ulcer (2, 3, 4, or unstageable) recorded during hospitalization according to National Pressure Injury Advisory Panel classifications.

### Histopathology analysis of tissue specimens

Histopathologic characteristics of biopsied patients were qualitatively and quantitatively assessed using hematoxylin & eosin (H&E) stained tissue specimens by a board-certified dermatopathologist (APF). For COVID-19(+) patients, 4mm biopsies from (typically) areas of sacral purpura were obtained. For control COVID-19(-) specimens, which were typically submitted to pathology during debridement, a search of Cleveland Clinic pathology archives was performed to find specimens from PUs. H&E sections of all available specimens were reviewed to identify those appropriate to use as control specimens. Any specimens with histologic features suggestive of secondary infection, extensive epidermal ulceration, or prominent fibrosis (supporting significant chronicity) were excluded. Ten specimens were found to have histologic features suggestive of inherent pressure-induced changes. For these, a 4mm wide area was demarcated and examined to be consistent with the area examined in COVID-19(+) tissue. When possible, areas with intact epidermis were chosen in COVID-19(-) specimens to also reduce variability with tissue examined from COVID-19(+) patients. Histopathologic features that were evaluated included neutrophil infiltrate, vascular thrombosis, total number of thromboses, intraluminal neutrophils and leukocytoclasis (**Table 3**). These features were scored for their presence or absence, in addition to quantitating them if present.

### NanoString analysis

RNA was extracted from Formalin-Fixed and Paraffin Embedded (FFPE) tissue biopsies, using the RNeasy FFPE kit (Qiagen) with deparaffinization solution (Qiagen). Total RNA concentrations were determined using a Nanodrop ND-1000 spectrophotometer (Thermo Fisher Scientific). One-hundred nanograms of RNA was then hybridized with the NanoString Human Autoimmune Profiling Panel and spike in Coronavirus Panel Plus probe sets (NanoString Technologies). Transcriptional analysis of immune related genes and SARS-CoV-2 genes in tissue biopsies was conducted using the NanoString nCounter Pro Analysis System (NanoString Technologies). Raw data was initially processed using the nSolver Analysis Software (V.4.0). Quality control and normalization was performed using the 6 positive control, 8 negative control, and 20 housekeeping probes. Samples flagged for failing quality control were removed. Remaining data was assessed for cell type scoring using nSolver Advanced Analysis (V.2.0) and exported for subsequent analysis in R software.

### Bioinformatics analysis

Analysis was performed using R (version 4.1.0). Venn diagram was generated utilizing DEGs from TV vs Control results against PU vs Control. Resulting residuals for principal component analysis (PC1 & PC2) were plotted using ‘ggplot2’ package (version 3.3.5). For pathway analysis, DEGs were separated into up- and down-regulated gene lists (fold change > 0 & fold change < 0 respectively). Package ‘gprofiler2’ (version 0.2.1) to find up- and down-regulated pathways. GO pathways were filtered to remove redundant pathways to generate the data by GO’s hierarchically structured database. Using ordered p-values from pathway analysis, the top 10 up-regulated and down-regulated pathways were obtained and p-values were log10-transformed for **Fig. 3A and Fig. 3B**. Heatmaps were generated using normalized counts for specific gene sets (pheatmap version 1.0.12). Gene set enrichment analysis (GSEA) was utilized to identify genes involved in specific biological functions like neutrophil activation, abnormal thrombosis and complement activation. COVID-19-specific neutrophil genes (**Fig. 4C**) were analyzed using published single-cell sequencing data. The p-value cutoffs that were used to determine significant genes from the NanoString data was FDR adjusted p-values < 0.05.

### Neutrophil extracellular traps (NETs) immunofluorescence staining and confocal imaging

Paraffin embedded tissue sections were sectioned (5 μm thickness) by the Cleveland Clinic Histology Core. Sections were deparaffinized and blocked using blocking buffer (Hank’s balanced salt solution containing 2% bovine serum albumin and 2% goat serum) for 1 hour at room temperature. Qualitative evaluation of presence of NETs in the tissue was performed using antibodies specific for NET staining including anti-neutrophil elastase (NE/ELA2; R&D Systems, Catalogue# MAB91671-100; monoclonal mouse IgG) and Anti-Histone H3 (citrulline R2 + R8 + R17; Abcam, Catalogue# ab5103; Rabbit polyclonal IgG). Primary antibody dilutions were made in the blocking buffer (NE, CitH3; used at 1:100 dilution) and added to the sections for an overnight incubation at 4 °C. Next day, slides were washed in 1X PBS (3X) and secondary antibodies were added to the sections for 1 hour at room temperature (1:1000 dilution in blocking buffer, Goat anti-Mouse 633 and Goat anti-Rabbit 568). After the 1 hour incubation, slides were washed in 1X PBS (3X) and mounted with DAPI (ProLong™ Gold Antifade Mountant with DAPI, ThermoFisher Scientific, Catalogue# P36931). Images were obtained using an inverted Leica SP8 confocal microscope using objective lens 40X oil/1X zoom factor. Five fields/tissue section were imaged for the purpose of quantitative assessment of presence of tissue NETs in the epidermis as well as the dermis. Images obtained from various disease groups were qualitatively assessed for presence of high, medium-low and no tissue NETs. Area of NETs was quantified using Image J software (version 1.53r 21).

### Statistical analysis

Categorical variables are presented as count (percentage) and continuous variables as median (interquartile range). Wilcoxon rank-sum was utilized for two-sample continuous variable comparisons. Chi-square and Fisher’s Exact tests were utilized to compare nominal categorical data across groups. Cochrane-Armitage exact trend test was utilized to compare univariate ordinal categorical data. Multivariable logistic and multivariable Cox proportional hazards regression were used to model binary and time-to-event outcomes, respectively. Multivariable linear regression was used to model wound size. Potential for multicollinearity was explored using variance inflation factor analysis. One-way ANOVA was utilized for analysis of tissue NETs with post-hoc Tukey analysis. An α level of 5% (two-sided) was utilized for all comparisons. Statistical analysis was performed using SAS JMP Pro (version 15) and Prism (version 9.3.1).

## Data Availability

All data produced in the present study are available upon reasonable request to the authors. The transcript profiling dataset will be deposited into a public database upon publication in a peer-reviewed journal.

## Acknowledgements

We would like to thank Emily Jiang from the Cleveland Clinic Department of Business Intelligence; Greg Strnad from the Cleveland Clinic COVID-19 Registry; and Janine Sot from the Cleveland Clinic Department of Dermatology.

## Funding

This publication was made possible by a COVID-19 Themed Pilot Grant Award (to APF and CM) from the Clinical and Translational Science Collaborative (CTSC) of Cleveland from the National Center for Advancing Translational Sciences (NCATS) component of the National Institutes of Health and NIH roadmap for Medical Research (UL1TR002548).

This work was also funded by the Office of the Assistant Secretary of Defense for Health Affairs through the Congressionally Directed Medical Research Programs Peer Reviewed Medical Research Program (W81XWH-16-0439/PR150299 to CM).

This study was also supported by the Crohn’s & Colitis Foundation (#662997 to SJ).

JN was supported by the Case Investigative & Translational Dermatology Training Program funded by the National Institute of Arthritis and Musculoskeletal and Skin Diseases of the National Institutes of Health (T32AR007569 to KC).

This work utilized the Leica SP8 confocal microscope that was purchased with funding from the National Institutes of Health Shared Instrumentation Grant 1S10OD019972-01. Opinions, interpretations, conclusions, and recommendations are those of the authors and are not necessarily endorsed by the funders.

## Author Contributions

Conceptualization: APF, JN, CM, SJ Methodology: APF, JN, CM, & SJ

Investigation: JN, SJ, AKP, RM, JG, AHW, SS, JJ, LK, ASN, JDM, EM-C, SG, KC, APF, CM

Visualization: JN, SJ, AKP, RM, JG, AHW, SS, JJ, LK, ASN, JDM, EM-C, SG, KC, APF, CM

Funding acquisition: APF, CM, JN, SJ, KC

Project administration: APF

Supervision: APF, CM Writing – original draft: JN, SJ

Writing – review & editing: JN, SJ, AKP, APF, CM

## Competing Interests

The authors have no conflicts to disclose.

## Data Availability

Nanostring transcript profiles will be publically available in the NCBI Gene Expression Omnibus. All other data are available in the main text or the supplementary materials.

## Supplementary Materials

**Supplementary Table 1**: Clinical and demographic characteristics of all patients hospitalized during the study period in relation to development of HASPI.

**Supplementary Table 2**: Comparison of hospital course in COVID-19 (+) patients with and without HASPIs.

**Supplementary Table 3**: Multivariable ordinal regression for worsening hospital course severity in COVID-19 (+) patients.

**Supplementary Table 4**: Thirty-day HASPI related morbidity.

**Supplementary Table 5**: ICD-10 codes used to search for healthcare acquired sacral pressure injury.

**Supplementary Figure 1:** Diagram showing the recruitment process.

**Supplementary Figure 2:** Diagram indicating sample sizes and molecular analysis parameters.

## Notes

### Competing Interest Statement

The authors have declared no competing interest.

### Author Declarations

The Institutional Review Board of Cleveland Clinic gave ethical approval for this work.

