## Supplementary Material for "Abnormal thrombosis and neutrophil activation increases the risk of hospital-acquired sacral pressure injuries and morbidity in patients with COVID-19"

**Supplementary Table 1:** Clinical and demographic characteristics of hospitalized patients during the study period.

| | | COVID (+) | COVID (-) | p-value[a] |
|---|---|---|---|---|
| Total Admissions | | 5,482 | 53,284 | |
| Total Hospitalization Days | | 64,980 | 526,783 | |
| Total HASPIs | | 49 | 244 | |
| HASPI Rate | | 7.5 per 10,000 days | 4.6 per 10,000 days | |
| Pressure Ulcer Risk at Admission[b] | | 19 (15-20) | 19 (16-21) | **<0.001** |
| Female Sex | | 2,635 (47.6%) | 26,620 (49.7%) | **0.003** |
| Age (Years) | | 69.4 (58.8-79.8) | 66.3 (53.5-76.7) | **<0.001** |
| Race | White | 3,513 (63.5%) | 37,962 (70.9%) | **<0.001** |
| | Black | 1,623 (29.3%) | 12,654 (23.6%) | |
| | Other | 395 (7.1%) | 2,912 (5.4%) | |
| Hospitalization Length (Days) | | 8 (6-14) | 7 (6-11) | **<0.001** |
| ICU Admission: Yes | | 2,210 (40.0%) | 18,656 (34.9%) | **<0.001** |
| Intubation: Yes | | 104 (1.9%) | 602 (1.1%) | **<0.001** |

**Bolded** values denote statistically significant p-values.

[a]Based on Wilcoxon-rank sum test for continuous variable comparisons and Chi-squared test for categorical variable comparisons.

[b]Based on Braden risk assessment score closest to admission. Note that a lower score equates to greater risk for ulcer formation.

**Supplementary Table 2**: Comparison of hospital course in COVID-19 (+) patients with and without HASPIs.

| | | HASPI (+) | HASPI (-) | p-value[a] |
|---|---|---|---|---|
| **N** | | **49** | **5,482** | **--** |
| Pressure Ulcer Risk at Admission[b] | | 14 (12-16.5) | 19 (15-20) | **<0.001** |
| Female Sex | | 17 (34.7%) | 2,618 (47.8%) | 0.08 |
| Age (Years) | | 66.3 (56.7-74.9) | 69.4 (58.9-79.8) | 0.22 |
| Race | White | 29 (59.2%) | 3,484 (63.6%) | 0.20 |
| | Black | 19 (38.8%) | 1,604 (29.3%) | |
| | Other | 1 (2.0%) | 394 (7.2%) | |
| Hospitalization Length (Days) | | 29 (19.5-35) | 8 (6-14) | **<0.001** |
| Co-morbidities | Hypertension | 31 (63.3%) | 3,230 (58.9%) | 0.53 |
| | COPD | 13 (26.5%) | 976 (17.8%) | 0.11 |
| | Diabetes | 23 (46.9%) | 1,912 (34.9%) | 0.08 |
| Hospital Course Severity | Hospitalization Only | 4 (8.2%) | 3,306 (60.3%) | **<0.001** |
| | ICU | 25 (51.0%) | 1,456 (26.6%) | |
| | Ventilation | 9 (18.4%) | 65 (1.2%) | |
| | Death | 11 (22.5%) | 655 (22.5%) | |

**Bolded** values denote statistically significant p-values.

[a]Based on Wilcoxon-rank sum test for continuous variable comparisons and Chi-squared or Fisher's Exact tests for categorical variables comparisons.

[b]Based on Braden risk assessment score closest to admission.

**Supplementary Table 3**: Multivariable ordinal regression for worsening hospital course severity in COVID-19 (+) patients.

| Outcome | Regression Estimate(95% CI) | p-value[a] |
|---|---|---|
| HASPI[b]: Yes vs. No | 2.2 (1.8-2.8) | **<0.001** |
| Age (Per additional year) | 0.99 (0.99-1.00) | 0.42 |
| Hypertension: Yes vs. No | 0.93 (0.88-0.99) | **0.01** |
| COPD[c]: Yes vs. No | 1.00 (0.99-1.1) | 0.11 |
| Diabetes: Yes vs. No | 1.1 (1.0-1.2) | **<0.001** |

**Bolded** values denote statistically significant p-values.

[a]These p-values represent multivariable ordinal regression analysis controlling for age and patient co-morbidities (hypertension, chronic obstructive pulmonary disease, congestive heart failure, and diabetes) with disease severity as the outcome.

[b]HASPI: Hospital-acquired sacral pressure injury.

[c]COPD: Chronic obstructive pulmonary disease.

**Supplementary Table 4**: Thirty-day HASPI-related morbidity.

| | COVID (+) | COVID (-) | p-value[a] |
|---|---|---|---|
| **N** | **47** | **247** | |
| 30-day morbidity[b] | 8 (16.3%) | 28 (11.5%) | 0.34 |
| Debridement: Yes | 7 (14.3%) | 14 (5.7%) | **0.047** |
| HASPI Infection: Yes | 4 (8.2%) | 20 (8.2%) | 0.99 |

**Bolded** values denote statistically significant p-values.

[a]Based on Fisher's Exact tests.

[b]Morbidity is defined as an aggregate of either requiring surgical debridement of HASPI or infection of HASPI from time of admission to 30-days after hospital discharge.

**Supplementary Table 5**: ICD-10 codes used to search for healthcare acquired sacral pressure injury.

| |
|---|
| L89.150, L89.151, L89.152, L89.153, L89.154, L89.159 - pressure ulcer of sacral region (various stages) |
| L98.429- non-pressure ulcer of sacrum |
| L98.499- non-pressure chronic ulcer of skin |
| L89.30, L89.31, L89.32, L89.302- pressure ulcer of buttocks |
| L98.411, L98.499- non-pressure ulcer of buttocks |
| L89.322, L89.323, L89.324, L89.312, L89.313, L89.314, L89.156, L89.90- pressure ulcer of left buttock (stages 2,3,4, or unspecified), right buttock (stages 2,3,4, or unspecified) |

**Supplementary Figure 1**

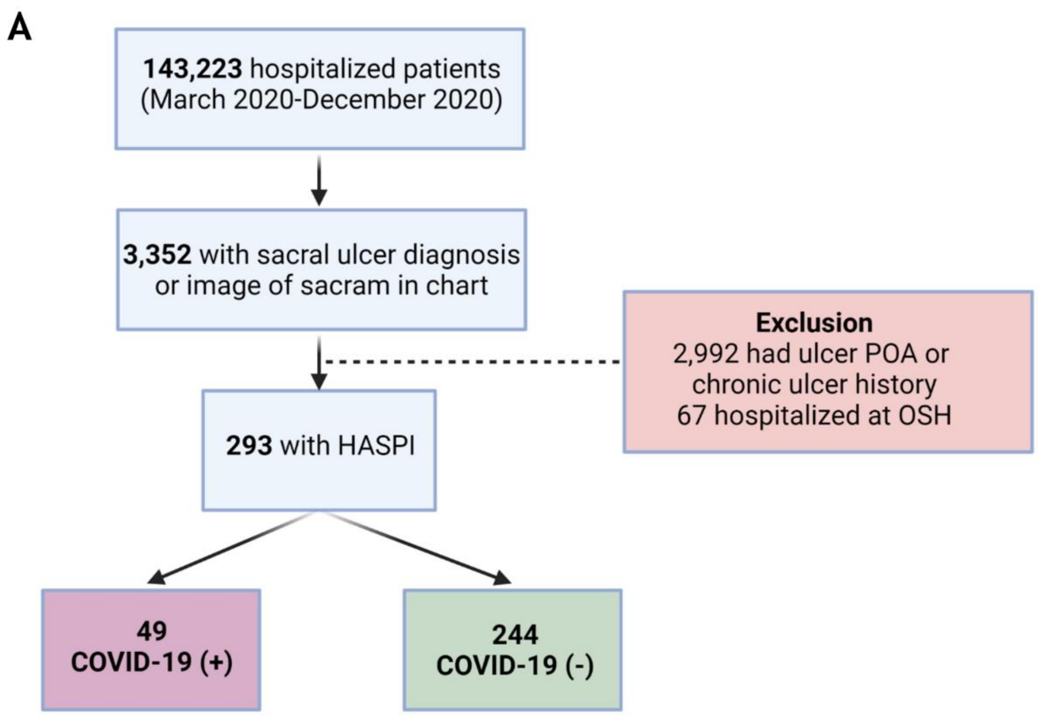


**Supplementary Fig. 1. Diagram showing the recruitment process.**

**(A)** Among the 143,527 patients hospitalized at our institution during our defined study period (March 2020-December 2020), 58,766 patients met our inclusion/exclusion criteria and comprised our study cohort. A total of 3,352 patients had a diagnosis of sacral ulcer during their hospitalization with or without an image of their sacral region in their electronic medical records. Of these patients, 2,992 had their sacral ulcers documented present on admission (POA) and 67 were seen at an outside hospital and were excluded from our study. Our final study cohort consisted of 293 patients with HASPIs in the COVID-19(+) and COVID-19(-) groups.

### Supplementary Figure 2

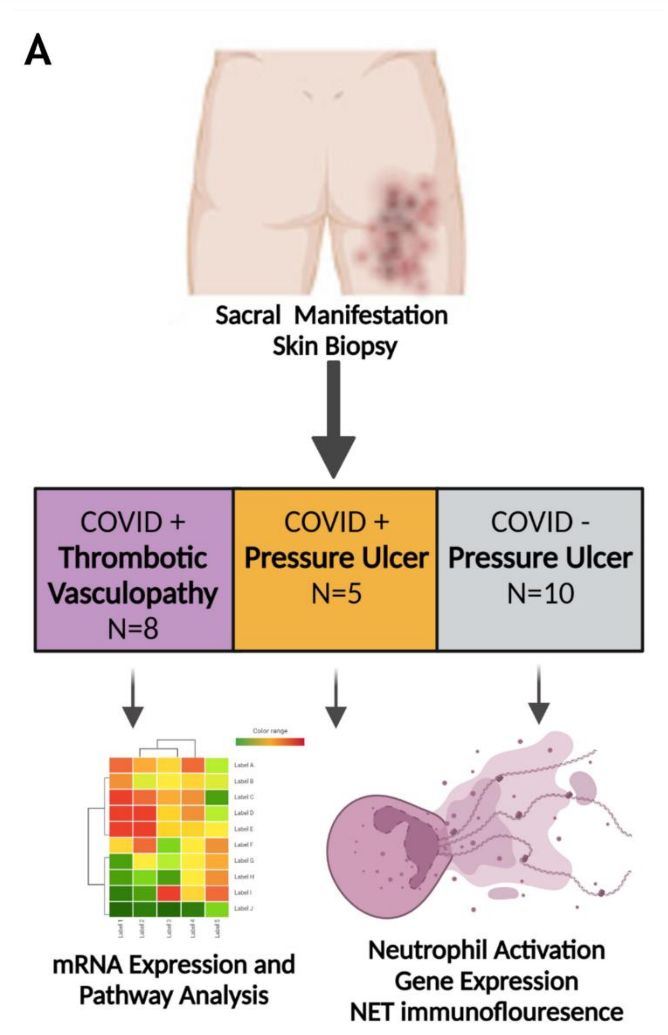


**Supplementary Fig. 2. Diagram indicating sample sizes and molecular analysis parameters.**

**(A)** Biopsies from COVID-19(+) patients with thrombotic vasculopathy (N=8), COVID-19(+) patients with pressure ulcers (N=5), and COVID-19(-) controls (N=10) were formalin fixed and paraffin embedded for histological analysis and staining to look at immune cell type specific markers. In addition, FFPE samples were utilized for NanoString analysis of mRNA expression in order to assess up- and down-regulated pathways as well as cell type scoring.
